# Efficacy of antimicrobial envelopes in preventing cardiac implantable electronic device infection – systematic review and meta-analysis

**DOI:** 10.1101/2025.03.14.25324008

**Authors:** Akanksha Mahajan, Ojas Mehta, Rhonda Stuart, Christopher Robson

## Abstract

Cardiac implantable electronic device (CIED) procedures have become increasingly common, accompanied by the challenge of CIED infections. This review aims to synthesise the available evidence to evaluate the efficacy of antibiotic eluting envelopes (AEEs) in preventing CIED infections and its effects on mortality.

All randomised controlled trials and observational studies that evaluated the efficacy of AEE use in reducing risk of CIED infections were included. Use of the TYRX AEE and CanGaroo envelopes hydrated in antibiotic solutions were considered for inclusion. The initial search yielded 493 articles, with 14 studies relevant for inclusion. A total of 87184 patients were included, with 14650 patients who received an AEE and 72534 patients who did not.

AEE use did not result in a statistically significant reduction in the odds of any CIED infection over total study duration (OR 0.73, 95% CI: 0.49-1.08), or within 12 months following CIED implantation (OR 0.85, 95% CI: 0.62-1.18). There was no reduction in odds of major CIED infection over total study duration (OR 0.73, 95% CI: 0.44-1.22) or within 12 months (OR 0.79, 95% CI: 0.46-1.37). The odds of minor CIED infection over any time (OR 0.75, 95% CI: 0.48-1.18) and overall mortality (OR 1.07, 95% CI: 0.60-1.88) were also not reduced.

However, subgroup analysis for patients at high risk of infection found that AEE use was associated with a reduction in total CIED infections over total study duration (OR 0.66, 95% CI: 0.45-0.97) and within 12 months (OR 0.73, 95% CI: 0.56-0.95).

## Introduction

Cardiac implantable electronic devices (CIED), including pacemakers, implantable cardioverter-defibrillators (ICDs), implantable cardiac monitors and cardiac resynchronization therapy (CRT) devices have revolutionised the field of cardiac electrophysiology through their ability to monitor and regulate cardiac rate and rhythm. These devices have both a diagnostic and therapeutic role, ranging from arrhythmia prevention and management to diagnostic tools for decision making on pacing ^1^.

CIED infections have become an emerging area of concern, as rates of CIED implantation have been increasing steadily worldwide ^2^. These infections hold great clinical significance, with substantial associated morbidity along with considerable healthcare costs, prolonged hospitalisations and repeat procedures ^3-5^. Recent studies assessing infection rates have found that overall CIED infection rates range from 1%-2% ^6-9^, with the greatest incidence in the initial 12 months after the CIED procedure and a gradual decline over time ^10, 11^; with a 2023 study reporting that the cumulative incidence of infection was 0.6%, 0.7%, and 0.9% within 3, 6, and 12 months, respectively ^12^. The organisms most commonly responsible for CIED infections are Staphylococcal species, especially *Staphylococcus aureus* and coagulase-negative staphylococci ^13, 14^; with methicillin-resistant *S. aureus* contributing significantly to CIED infection related mortality ^14^.

There are several established risk factors for CIED infection, including patient factors such as sex, age, and comorbidities, as well as procedure-related factors such as type of device, re-operation, temporary pacing and presence of epicardial leads ^15-17^. However, these factors are mostly non-modifiable, presenting a need for more effective infection prevention protocols, beyond the current standard-of-care measures such as chlorhexidine skin preparation and peri-procedural systemic antimicrobial prophylaxis ^1^. Insertion of an antibiotic-eluting envelope (AEE) at the time of device implantation to prevent early infection has been investigated. Until recently, the only product approved for clinical use was the TYRX (Medtronic) AEE, which consists of a multifilament mesh envelope that delivers a sustained combination of rifampicin and minocycline into the local tissue over 7 days, following which the mesh fully absorbs in 9 weeks ^18^. Recently, EluPro (formerly CanGaroo Envelope) (Aziyo Biologics, Silver Spring, MD), a drug-eluting envelope also containing rifampicin and minocycline delivered over a target duration of 7 days, constructed from reinforced layers of natural extracellular tissue matrix, has received United States Food and Drug Administration (FDA) clearance in 2024 ^19, 20^.

Although the efficacy of AEEs has been evaluated in previous reviews, there have been several studies published in recent years. Furthermore, recent randomised controlled trials and observational studies have demonstrated conflicting results. This systematic review and meta-analysis aims to synthesise the available evidence to provide an updated evaluation of the efficacy of AEEs in preventing CIED infections in patients undergoing CIED implantation or upgrade procedures, as well as its effects on mortality and length of hospitalisation.

## Methods

This review protocol was registered with the International Register of Prospective Systematic Reviews (PROSPERO) under the CRD number CRD42024588950. This review was conducted according to the Preferred Reporting Items for Systematic Review and Meta-Analysis (PRISMA) guidelines.

### Eligibility Criteria

All human randomised controlled trials and observational studies (case-control and cohort) that evaluated the efficacy of use of AEEs in reducing risk of CIED infections compared to no AEE use were included. Patients in any healthcare setting were considered. Use of the TYRX AEE, as well as CanGaroo biologic envelopes which were hydrated in antibiotic solutions were considered for inclusion. However, only studies which evaluated the TYRX envelope were included in the meta-analysis. Animal and in-vitro studies were excluded, as were studies where both the intervention and control group received an AEE.

### Search strategy

The literature was systematically searched from inception until September 2024 to identify relevant studies from a range of databases including MEDLINE, CINAHL, Embase, Scopus and Cochrane Library. An English translation was obtained for any articles not published in English. The following key words were used: “antimicrobial envelope”, “antibiotic envelope”, “TYRX”, “AIGISRx”, “pacemaker,” “cardiac resynchronization therapy devices”, “defibrillators”. This search was supplemented with a manual search of the reference lists of relevant articles as well as grey literature such as conference abstracts and posters.

### Data extraction

Search data were uploaded to the Covidence review platform (Covidence, Australia), and duplicates were removed. Two reviewers (AM and CR) independently screened the title and abstract of the identified citations. Full texts were subsequently independently screened by two reviewers (AM and CR) for eligibility. Disagreements were resolved by discussion among the authors with a consensus decision being reached. Data was extracted from eligible studies including year of publication, study design, sample size, definition of exposure and outcome, inclusion and exclusion criteria, adjusted covariates, measures of the association including odds ratio and relative risk. Outcome data related to overall infection rates, major infection rates, local/minor infection rates, and mortality were collected. Major CIED infection was defined as infection with positive blood cultures, requiring systemic antibiotics, infective endocarditis, bacteraemia, or repeat surgical intervention (e.g. device explantation). Local/minor CIED infections were defined as those which did not meet the criteria for major infection. Patients at high risk of CIED infection were defined as those undergoing CIED generator replacement or a system upgrade, CIED pocket or lead revision, or an initial CRT-D procedure, as per the WRAP-IT trial ^7^.

### Quality assessment

The RoB 2 tool for randomised and ROBINS-I tool for non-randomised trials was used to assess the quality and risk of bias in all included studies. The RoB 2 tool involves scoring studies as ‘low risk’, ‘some concerns’ or ‘high risk’ across 5 domains of bias, based on responses to a series of signalling questions in each domain ^21^. The ROBINS-I tool involves scoring studies across 7 domains of bias, with 2 pre-intervention domains, 1 at-intervention, and 4 domains which assess the risk of bias arising post-intervention ^22^. Risk-of-bias plots were generated using the robvis tool ^23^.

### Statistical analysis

Meta-analysis of Observational Studies in Epidemiology (MOOSE) guidelines were followed to conduct the meta-analysis. All statistical analyses were performed using Review Manager (RevMan) version 7.2.0. Forest plots were generated to demonstrate the effect of each study and the pooled effect size for studies with the same outcome. The probability value of p < 0.05 was considered statistically significant. A random effects model was used for meta-analysis as it accounts for heterogeneity between studies and allows for generalisation of results beyond the individual studies included in the meta-analysis ^24^. The effects of measure used for dichotomous outcomes related to CIED infections and overall mortality were unadjusted odds ratio (OR) along with their corresponding 95% CI. The unadjusted OR was calculated by the authors when it was not included in the published study, but adequate data was provided for calculation.

For each overall effect size, heterogeneity was examined using Cochran’s Q statistic (measure of weighted square deviations), with N-1 degrees of freedom (where N is the number of studies), between studies variance (T2), and ratio of the true heterogeneity to total observed variation (I^2^). The I^2^ values were used to quantify heterogeneity using the following parameters: 0% no heterogeneity, 25% low heterogeneity, 50% moderate heterogeneity, and 75% high heterogeneity ^25^. Potential causes for heterogeneity were evaluated through sensitivity analysis. Publication bias was assessed through visual inspection of funnel plots.

## Results

### Search results and study characteristics

The initial search yielded 493 articles. After removal of duplicates, the title and abstracts of 332 articles were reviewed by 2 authors (AM and CR). The full texts of 33 articles were reviewed by AM and CR, with 14 studies deemed relevant for inclusion, including 13 observational studies ^26-38^, with 8 retrospective cohort studies ^26, 27, 29, 30, 33-36^, 4 prospective cohort studies ^31, 32, 37, 38^, 1 cohort study which combined retrospective and prospective data ^28^, as well as 1 randomised controlled trial ^7^. Nine studies were excluded due to a lack of appropriate comparator/control group, and 1 study was excluded because it assessed geographical variations in patient outcomes after CIED implantation rather than efficacy of AEEs. A further 9 studies were excluded because they presented results from a patient cohort which had also been published in another included article. For such studies, the article that was published most recently, and did not analyse a specific subgroup of the total sample population was included. The PRISMA flow diagram is shown in Figure 1.

**Figure 1:**
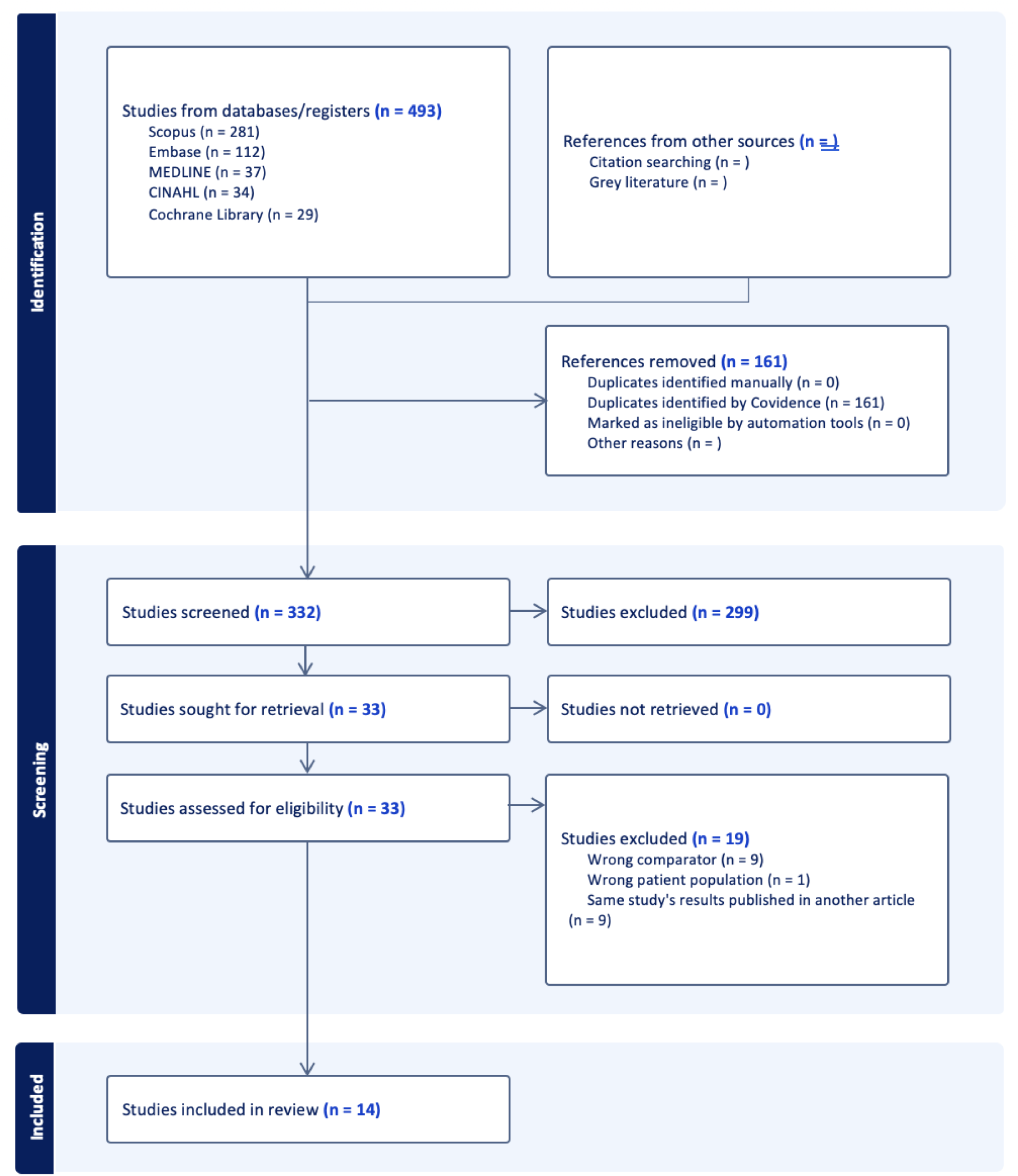
PRISMA Flowchart.

A total of 87184 patients were included, with 14650 patients who received an AEE and 72534 patients who did not. There were 8 studies from the United States of America, 2 from Italy ^32, 38^, as well as 1 study each from Sweden ^27^ and Denmark ^29^; with all studies being conducted in tertiary hospitals. All studies reported on the use of the TYRX (AIGISRx) AEE, except for one study which reported results from the SECURE and CARE observational studies which examined efficacy of the CanGaroo Envelope (Aziyo Biologics, Silver Spring, MD), when soaked in antibiotic solutions. The study characteristics are detailed in Table 1, and inclusion and exclusion criteria for each study in Supplementary Table S1. The definition of CIED infection, major and local infection outcomes in each study is provided in Supplementary Table S2. The definition of ‘high risk’ patient groups in included studies varied across studies and is outlined in Supplementary Table S3.

**Table 1:** Study characteristics of included studies.

| Study ID | Location | Time period | Study Design | Duration of follow up | Total sample size | Mean age (years) ± SD | Gender (%male) | Number of participants who received AEE | Number of Controls |
| --- | --- | --- | --- | --- | --- | --- | --- | --- | --- |
| Asundi 2019 | United States of America | 2008-2015 | Retrospective cohort | 6 months | 2059 | median 71.7 | 97.9% | 122 | 1976 |
| Chaudhry 2022 | Sweden | 2017-2019 | Retrospective cohort | 12 months | 526 | 72 | 69% | 144 | 382 |
| Deering 2022 | United States of America | 2014-2017 | | mean 223.7 ± 173.0 days | 1102 | 72.0 ± 12.0 | 60.9% | 850 | 252 |
| Frausing 2022 | Denmark | 2008-2019 | Retrospective cohort | 24 months | 1943 | 72.6 | 80% | 736 | 1207 |
| Hassoun 2017 | United States of America | 2014 | Retrospective cohort | Mean follow up 9 months in both groups | 184 | 71 | 60% | 92 | 92 |
| Henrikson<br>2017 | United States<br>of America | 2009-2013 | Prospective cohort | 12 months | 1707 | N/A | 76% | 1129 | 578 |
| Imberti<br>2023 | Italy | 2016-2020 | Prospective cohort | median follow-up of<br>42.3 (30.2–56.4)<br>months | 838 | median 77 | 65.4% | 46 | 792 |
| Kolek<br>2015 | United States<br>of America | 2009-2014 | Retrospective cohort | Minimum 300 days<br>(9.86 months) | 1124 | 69 | 34.7% | 488 | 636 |
| Mittal<br>2014 | United States<br>of America | 2007-2011 | Retrospective cohort | 6 months | 2880 | 77 | 62% | 275 | 2616 |
| Shariff<br>2015 | United States<br>of America | 2011-2013 | Retrospective cohort<br>study | 6 months | 1476 | 68 | 65% | 365 | 1111 |
| Tarakji<br>2019 | United States<br>of America | 2015-2017 | Randomized<br>controlled trial | 36 months<br>(mean 20.7±8.5<br>months) | 6983 | 70.1±12.5 | 71.7% | 3495 | 3488 |
| Shatla<br>2024 | United States<br>of America | 2016-2019 | Retrospective cohort | 6 months | 64295 | 72.7±12.9 | 58% | 5845 | 58450 |
| Woodard<br>2022 | United States<br>of America | 2017-2019 | Prospective cohort | 12 months | 248 | 71.6 ± 13.3 | - | 191 | 57 |
| Ziacchi<br>2023 | Italy | 2016-2022 | Prospective cohort | 5 years | 1819 | 72.8 ± 14.0 | 69.5% | 872 | 947 |

### Quality Assessment

There was variation in the methodological quality of included studies as summarised in Figure 2. Of the 13 observational studies included, 8 had low risk of bias, 3 had moderate risk of bias, and 2 had serious risk. The randomised controlled trial included was of high methodological quality and had low risk of bias.

**Figure 2:**
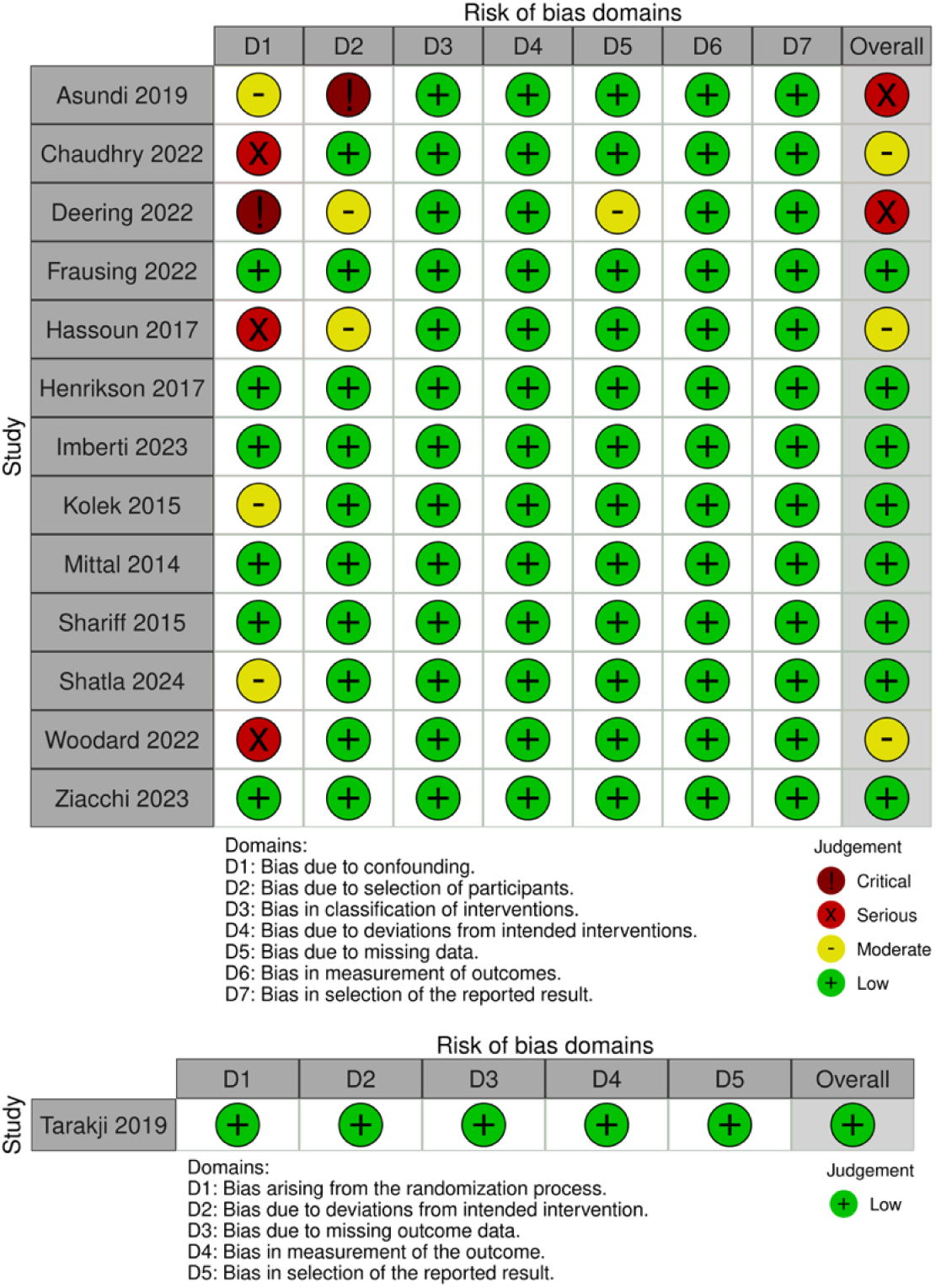
Methodological quality of included observational studies as per the ROBINS I tool (top) and included randomised study as per the RoB 2 tool (bottom)

### Meta analysis

The study by Woodard (2022) ^37^ was excluded from the meta-analyses for an association between AEE use and CIED infections because no CIED infections occurred in both the group that received the TYRX AEE as well as the controls, making the effect size inestimable. The study by Deering (2022) ^28^ was excluded because the patients in the intervention arm received the CanGaroo envelope, as opposed to the TYRX envelope which was used in all other studies. This study was also excluded because several different antibiotic combinations were used to hydrate the envelope, including gentamicin; whereas all studies included in the meta-analysis used only a combination of rifampicin and minocycline. The study by Henrikson et al ^31^ reported on a combination of results from the Centurion (CRT recipients) and Citadel (ICD recipients) studies. Results for site matched, comorbidity matched and previously published controls were provided for the Citadel study, whereas only previously published controls were used as a comparison group for the Centurion study. Therefore, the site matched and comorbidity matched control groups from the Centurion study were considered in this meta-analysis; while the Citadel study was excluded due to the lack of a true control group.

This review found that AEE use did not result in a statistically significant reduction in the odds of any type of CIED infection over total study duration, as shown in Figure 3 (OR 0.73, 95% CI: 0.49-1.08, p=0.12), or within 12 months following CIED implantation, as demonstrated in Supplementary Figure S1. Moreover, there was no statistically significant reduction in the odds of major CIED infection over total study duration or within 12 months as demonstrated in Supplementary Figure S2 and S3 respectively. The odds of minor CIED infection over any time and overall mortality were also not statistically significantly reduced in the AEE group, as shown in Supplementary Figure S4 and S5.

**Figure 3:**
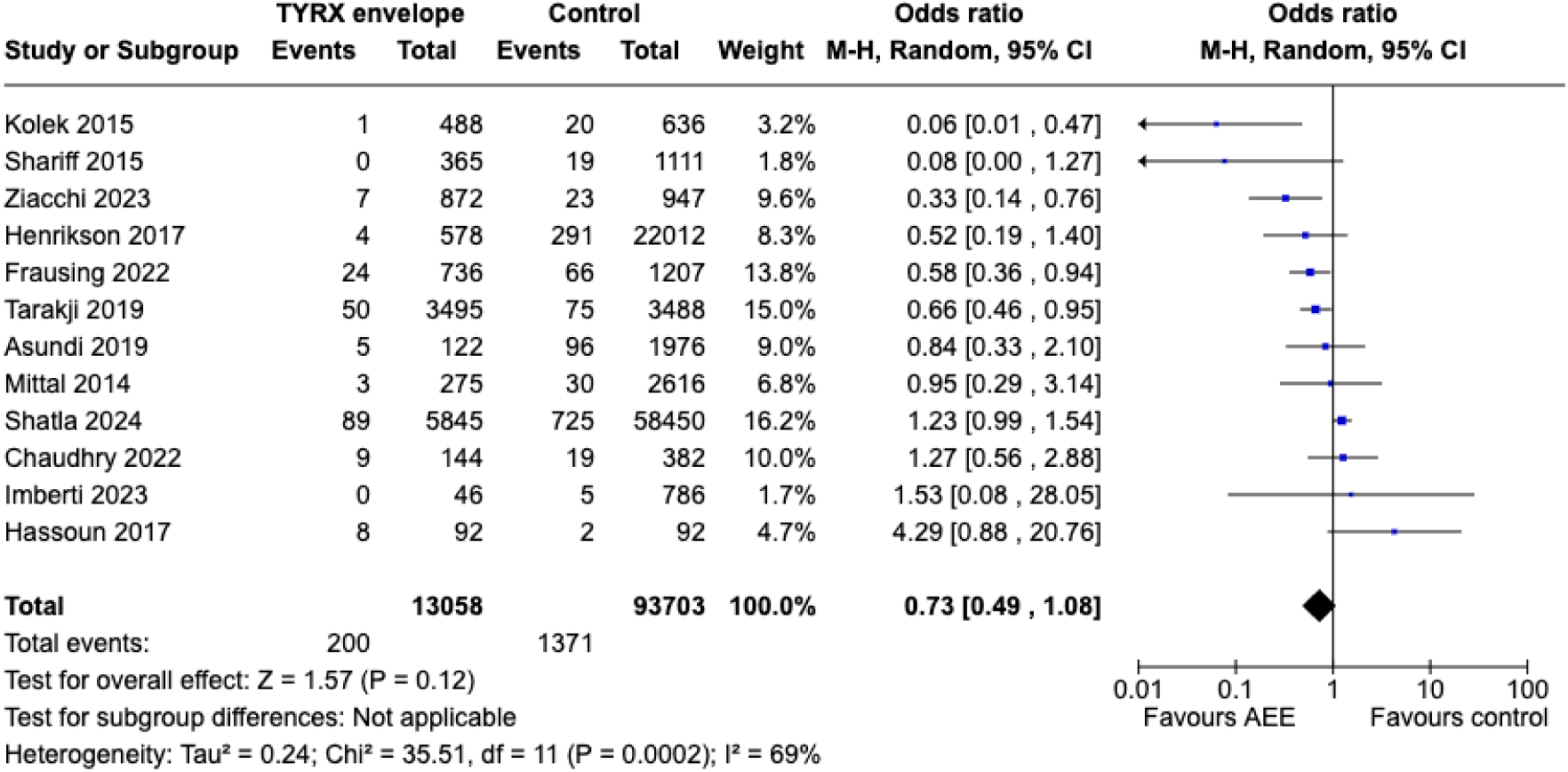
Forest plot depicting the pooled odds ratio for any CIED infection over entire study period in AEE vs control groups.

However, sensitivity analysis which excluded studies at moderate, serious or critical risk of bias ^26, 27, 30^ demonstrated that AEE use was associated a statistically significant reduction in total CIED infections (OR 0.59, 95% CI: 0.37-0.95, p= 0.03) and major CIED infections (OR 0.55, 95% CI 0.41-0.74, p<0.01) over total study duration, as well as major CIED infections within 12 months (OR 0.62, 95% CI: 0.44-0.87, p<0.01), as shown in Figure 4 and Supplementary Figure S6. Additionally, as demonstrated in Figure 5 and Supplementary Figure S7, subgroup analysis for studies which evaluated the efficacy of AEEs in patients at high risk of infection found that AEE use was associated with a statistically significant reduction in total CIED infections over total study duration (OR 0.66, 95% CI: 0.45-0.97, p=0.03) as well as within 12 months after the procedure (OR 0.73, 95% CI: 0.56-0.95, p=0.02). Nevertheless, AEE use was not associated with reduced risk of local/minor infections or major CIED infections over total study duration and did not reduce mortality in high risk groups.

**Figure 4:**
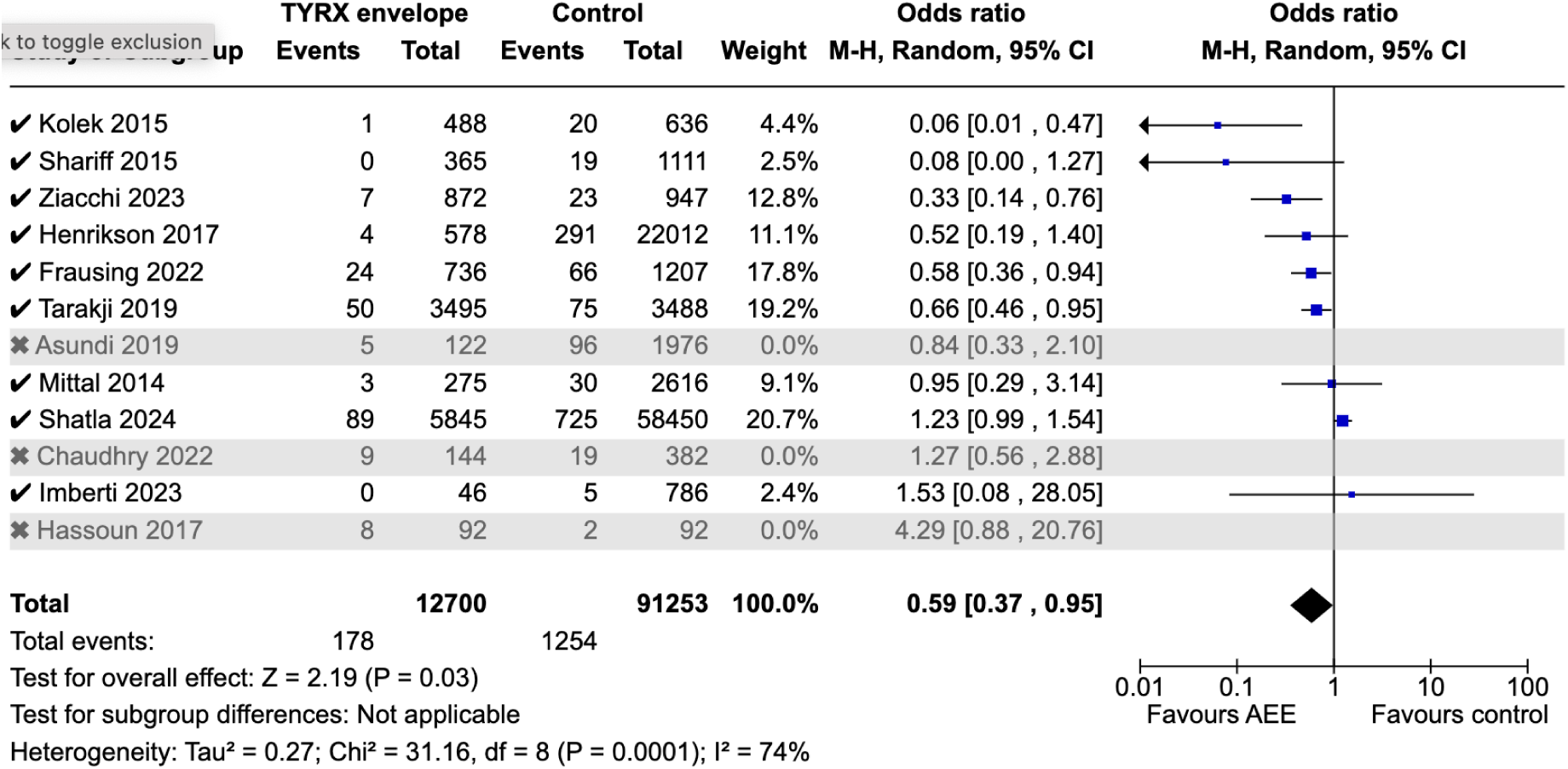
Sensitivity analysis for total CIED infections over total study duration, including studies at low risk of bias only; excluding studies at moderate, serious or critical risk of bias.

**Figure 5:**
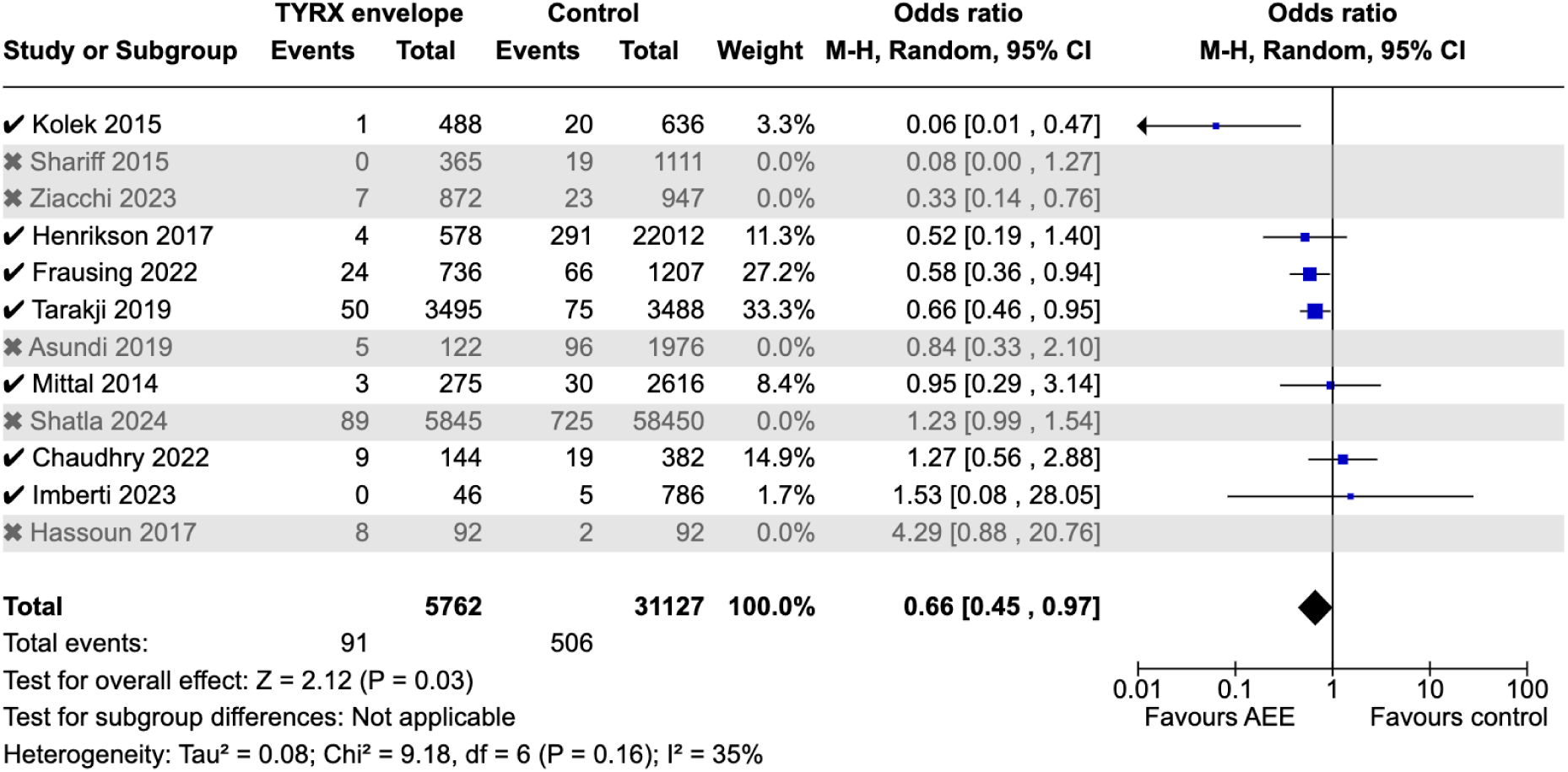
Sensitivity analysis for total CIED infections over total study duration including studies which assigned AEE to high risk groups only.

The funnel plot for all studies included in this review has a symmetrical distribution, suggesting that there is low publication bias (Supplementary Figure S8). Additionally, there is moderate heterogeneity in the data overall for all included studies in this meta-analysis, based on the analysis for any CIED infection over the entire study period (I^2^ value 69% in Figure 3). However, there was low heterogeneity in the data in studies reporting on minor CIED infections (I^2^ 2% in Supplementary Figure S4).

## Discussion

This is the first systematic review to examine the impact of AEE on all CIED infections, including local infections; as previous reviews have only examined the outcomes of major CIED infection and mortality. It is also the first to conduct subgroup analysis for infection risk within 12 months and compare it to entire study periods which are of varying durations. Previous meta analyses published in 2020 ^39-41^ reported a statistically significant reduction in major CIED infection with the use of AEE. Our review found no statistically significant reduction when all available studies were included. Nevertheless, the associations found in this review’s sensitivity analysis including only studies at low risk of bias, as well as the subgroup analysis evaluating efficacy of AEEs for reducing total CIED infections in high risk groups aligns with the findings of previous reviews. Ullah et al ^40^, Asbeutah et al ^39^ and Pranata et al ^41^ each included only the same 6 studies in both the qualitative synthesis and meta-analysis and acknowledge the likely presence of publication bias due to the limited number of studies included. However, our systematic review includes 14 studies in the qualitative synthesis and 12 studies in the meta-analysis and demonstrates lower publication bias based on the funnel plot compared to previous reviews.

There are a number of potential sources of bias which may have influenced the results of those studies which found that AEEs were not effective in reducing odds of CIED infection, and in fact suggested that they may increase this risk. In the paper by Hassoun et al ^30^, although patients with AEE had higher rates of CIED infections, they also had a greater number of risk factors for infection at baseline, which is a significant confounding factor and likely accounts for their results which contradict that of other studies. Similarly, the envelope group had a higher total PADIT score ^42^ at baseline compared to the control group in the study by Chaudhry et al ^27^; with significant differences for the subcomponents: depressed renal function, immunosuppression, and prior procedures on the same pocket. This may have negatively skewed the results related to AEE efficacy. Although there were no significant differences in baseline parameters between groups in the study by Imberti et al ^32^, there were lower number of events of CIED infection (0.6%) compared to the other studies, which may increase risk of bias in the results. Moreover, the results of this study may differ from other included studies due to use of AEEs only in ICD/CRT patients deemed at high risk of infection according to a PADIT score > 6 or dialysis; with no pacemaker patients receiving AEE. ICD and CRT procedures inherently present a higher risk of infection^8, 16^.

Antibiotic envelope use has significant associated costs, and the cost-efficacy of this intervention must be considered, especially given the potentially limited efficacy in infection prevention. There are several studies which agree that use of the TYRX envelope has favourable cost-effectiveness ratios below accepted willingness-to-pay thresholds for patients at high risk of post-operative infection, resulting in lower overall healthcare resource utilization despite the initial cost ^43-46^. However, some studies report that an AEE may not be economically favourable in patients with low infection risk ^44, 45, 47^. This is particularly important when considering the integration of such an intervention into public healthcare system guidelines. Moreover, many included studies included AEE insertion in patient populations that had higher infection risk compared to the control group, suggesting that these results may not be applicable to all patient populations.

The antimicrobial stewardship implications of this intervention must also be taken into account when considering its utility. Given that AEEs are impregnated with rifampicin and minocycline, this antimicrobial exposure on a large scale has the potential to contribute to antimicrobial resistance at an individual and population level. Rifampicin resistance in particular is a growing public health concern as it can develop rapidly following mutations in the rpoB gene encoding the β-subunit of bacterial DNA-dependent RNA polymerase ^48, 49^. There appears to be a growing tendency in antibiotic-resistant strains of staphylococci ^50, 51^ which is associated with higher infection recurrence rates by enhancing biofilm formation and reducing susceptibility to vancomycin ^52^. Nevertheless, a systematic review ^53^ of 44 studies evaluating the potential for developing new antimicrobial resistance from the use of antimicrobial medical devices has reported that although new antibiotic resistance was commonly reported with the use of rifampicin as a single agent; it was only reported in two^54, 55^ out of nine studies assessing the minocycline/rifampicin combination. Further longitudinal studies are required in this area to clarify the risk of antimicrobial resistance and its impacts.

### Limitations

There were significant group differences in some of the included studies ^28-30, 34, 37, 38^, with the AEE group having higher pre-procedure infection risk compared to the control group. This may have skewed the results to suggest that the AEE was ineffective due to greater infection rates which may actually be attributable to other confounding factors. Therefore, further observational studies and randomised controlled trials which include patients that are at similar risk of infection, or observational studies with propensity-matched AEE and control groups are required to validate the findings of this review. Additionally, although some included studies ^26, 32^ also reported infection rates in groups with comorbidities such as diabetes, cerebrovascular diseases and chronic obstructive pulmonary disease separately, this data was not reported in most included studies and further studies are required to elucidate the potential varying impact of different comorbidities in infection risk and hence indication for AEE use.

## Conclusion

Based on the available data, AEEs are associated with lower odds of major CIED infection in high risk patients undergoing CIED procedures; however, they do not reduce risk of minor infection or overall mortality. While there are certain populations at higher risk of infection where CIED AEEs have been shown to provide clinical benefit, patient selection remains an important part of the process. Additionally, further research is required to explore the CIED infection risk associated with different patient comorbidities and hence refine the definition of ‘high risk’ patients for CIED infection, in order to develop clear guidelines for indications for AEE use for CIED infection prophylaxis.

The majority of patients that undergo CIED procedures are not part of the high-risk populations in which AEEs are likely most cost-effective. Moreover, reduction in infection risk and subsequent savings in healthcare costs to institutions, governments and individuals needs to be justified against the cost of AEEs themselves. Therefore, in the context of increasing antimicrobial resistance, judicious use of the AEEs is an important duty of the implanting physician and further evidence regarding AEE efficacy is required to inform guidelines for their use to reduce unnecessary healthcare expenditure.

## Data Availability

All data supporting the findings of this study are available within the paper and its Supplementary Materials.

## Acknowledgements

No funding was obtained for this publication.

## References

1. Glikson M, Nielsen JC, Kronborg MB, Michowitz Y, Auricchio A, Barbash IM, et al. 2021 esc guidelines on cardiac pacing and cardiac resynchronization therapy: Developed by the task force on cardiac pacing and cardiac resynchronization therapy of the european society of cardiology (esc) with the special contribution of the european heart rhythm association (ehra). European Heart Journal. 2021;42:3427–3520

2. Han HC, Hawkins NM, Pearman CM, Birnie DH, Krahn AD. Epidemiology of cardiac implantable electronic device infections: Incidence and risk factors. Europace. 2021;23:iv3-iv10

3. Greenspon AJ, Eby EL, Petrilla AA, Sohail MR. Treatment patterns, costs, and mortality among medicare beneficiaries with cied infection. Pacing and Clinical Electrophysiology. 2018;41:495–503

4. Wilkoff BL, Boriani G, Mittal S, Poole JE, Kennergren C, Corey GR, et al. Impact of cardiac implantable electronic device infection: A clinical and economic analysis of the wrap-it trial. Circ Arrhythm Electrophysiol. 2020;13:e008280

5. Joy PS, Kumar G, Poole JE, London B, Olshansky B. Cardiac implantable electronic device infections: Who is at greatest risk? Heart Rhythm. 2017;14:839–845

6. Birnie DH, Wang J, Alings M, Philippon F, Parkash R, Manlucu J, et al. Risk factors for infections involving cardiac implanted electronic devices. Journal of the American College of Cardiology. 2019;74:2845–2854

7. Tarakji KG, Mittal S, Kennergren C, Corey R, Poole JE, Schloss E, et al. Antibacterial envelope to prevent cardiac implantable device infection. N Engl J Med. 2019;380:1895–1905

8. Yang PS, Jeong J, You SJ, Yu HT, Kim TH, Sung JH, et al. The burden and risk factors for infection of transvenous cardiovascular implantable electronic device: A nationwide cohort study. Korean Circ J. 2019;49:742–752

9. Clémenty N, Carion PL, Léotoing L, Lamarsalle L, Wilquin-Bequet F, Brown B, et al. Infections and associated costs following cardiovascular implantable electronic device implantations: A nationwide cohort study. Europace. 2018;20:1974–1980

10. Johansen JB, Jørgensen OD, Møller M, Arnsbo P, Mortensen PT, Nielsen JC. Infection after pacemaker implantation: Infection rates and risk factors associated with infection in a population-based cohort study of 46299 consecutive patients. Eur Heart J. 2011;32:991–998

11. Uslan DZ, Sohail MR, St Sauver JL, Friedman PA, Hayes DL, Stoner SM, et al. Permanent pacemaker and implantable cardioverter defibrillator infection: A population-based study. Arch Intern Med. 2007;167:669–675

12. Han H-C, Wang J, Birnie DH, Alings M, Philippon F, Parkash R, et al. Association of the timing and extent of cardiac implantable electronic device infections with mortality. JAMA Cardiology. 2023;8:484–491

13. Hussein AA, Baghdy Y, Wazni OM, Brunner MP, Kabbach G, Shao M, et al. Microbiology of cardiac implantable electronic device infections. JACC: Clinical Electrophysiology. 2016;2:498–505

14. Greenspon AJ, Rhim ES, Mark G, Desimone J, Ho RT. Lead-associated endocarditis: The important role of methicillin-resistant staphylococcus aureus. Pacing Clin Electrophysiol. 2008;31:548–553

15. Olsen T, Jørgensen OD, Nielsen JC, Thøgersen AM, Philbert BT, Frausing M, et al. Risk factors for cardiac implantable electronic device infections: A nationwide danish study. Eur Heart J. 2022;43:4946–4956

16. Olsen T, Jørgensen OD, Nielsen JC, Thøgersen AM, Philbert BT, Johansen JB. Incidence of device-related infection in 97 750 patients: Clinical data from the complete danish device-cohort (1982-2018). Eur Heart J. 2019;40:1862–1869

17. Sohail MR, Hussain S, Le KY, Dib C, Lohse CM, Friedman PA, et al. Risk factors associated with early-versus late-onset implantable cardioverter-defibrillator infections. Journal of Interventional Cardiac Electrophysiology. 2011;31:171–183

18. Hansen LK, Brown M, Johnson D, Palme Ii DF, Love C, Darouiche R. In vivo model of human pathogen infection and demonstration of efficacy by an antimicrobial pouch for pacing devices. Pacing and Clinical Electrophysiology. 2009;32:898–907

19. Elutia announces fda clearance of elupro™: The first antibiotic-eluting bioenvelope designed to protect patients with implantable cardiac pacemakers and defibrillators. 2024

20. Garrigos ZE, Catanzaro JN, Deegan D, Zhang J, Sohail MR. Preclinical evaluation of a novel antibiotic-eluting bioenvelope for cied infection prevention. Frontiers in Drug Delivery. 2024;4:1441956

21. Higgins JPT, Savović J, Page MJ, Elbers RG, Sterne JAC. Assessing risk of bias in a randomized trial. Cochrane handbook for systematic reviews of interventions. 2019:205–228.

22. Sterne JA, Hernán MA, Reeves BC, Savović J, Berkman ND, Viswanathan M, et al. Robins-i: A tool for assessing risk of bias in non-randomised studies of interventions. Bmj. 2016;355:i4919

23. McGuinness LA, Higgins JPT. Risk-of-bias visualization (robvis): An r package and shiny web app for visualizing risk-of-bias assessments. Research Synthesis Methods. 2020;n/a

24. Field AP, Gillett R. How to do a meta-analysis. Br J Math Stat Psychol. 2010;63:665–694

25. Khoshdel A, Attia J, Carney S. Basic concepts in meta-analysis: A primer for clinicians. International journal of clinical practice. 2006;60:1287–1294

26. Asundi A, Stanislawski M, Mehta P, Baron AE, Mull HJ, Ho PM, et al. Real-world effectiveness of infection prevention interventions for reducing procedure-related cardiac device infections: Insights from the veterans affairs clinical assessment reporting and tracking program. Infection Control and Hospital Epidemiology. 2019;40:855–862

27. Chaudhry U, Borgquist R, Smith JG, Mortsell D. Efficacy of the antibacterial envelope to prevent cardiac implantable electronic device infection in a high-risk population. Europace : European pacing, arrhythmias, and cardiac electrophysiology : journal of the working groups on cardiac pacing, arrhythmias, and cardiac cellular electrophysiology of the European Society of Cardiology. 2022;24:1973–1980

28. Deering TF, Catanzaro JN, Woodard DA. Physician antibiotic hydration preferences for biologic antibacterial envelopes during cardiac implantable device procedures. Frontiers in Cardiovascular Medicine. 2022;9

29. Frausing MHJP, Nielsen JC, Johansen JB, Jorgensen OD, Gerdes C, Olsen T, et al. Rate of device-related infections using an antibacterial envelope in patients undergoing cardiac resynchronization therapy reoperations. Europace. 2022;24:421–429

30. Hassoun A, Thottacherry ED, Raja M, Scully M, Azarbal A. Retrospective comparative analysis of cardiovascular implantable electronic device infections with and without the use of antibacterial envelopes. The Journal of hospital infection. 2017;95:286–291

31. Henrikson CA, Sohail MR, Acosta H, Johnson EE, Rosenthal L, Pachulski R, et al. Antibacterial envelope is associated with low infection rates after implantable cardioverter-defibrillator and cardiac resynchronization therapy device replacement: Results of the citadel and centurion studies. JACC. Clinical electrophysiology. 2017;3:1158–1167

32. Imberti JF, Mei DA, Fontanesi R, Gerra L, Bonini N, Vitolo M, et al. Low occurrence of infections and death in a real-world cohort of patients with cardiac implantable electronic devices. Journal of Clinical Medicine. 2023;12

33. Kolek MJ, Patel NJ, Clair WK, Whalen SP, Rottman JN, Kanagasundram A, et al. Efficacy of a bio-absorbable antibacterial envelope to prevent cardiac implantable electronic device infections in high-risk subjects. Journal of cardiovascular electrophysiology. 2015;26:1111–1116

34. Mittal S, Shaw RE, Michel K, Palekar R, Arshad A, Musat D, et al. Cardiac implantable electronic device infections: Incidence, risk factors, and the effect of the aigisrx antibacterial envelope. Heart rhythm. 2014;11:595–601

35. Shariff N, Eby E, Adelstein E, Jain S, Shalaby A, Saba S, et al. Health and economic outcomes associated with use of an antimicrobial envelope as a standard of care for cardiac implantable electronic device implantation. Journal of cardiovascular electrophysiology. 2015;26:783–789

36. Shatla I, Mehta NA, Kennedy KF, Elkaryoni A, Wimmer AP. Antibacterial envelope use to prevent cardiac implantable device infection: Outcomes from the national readmission database. Journal of Interventional Cardiac Electrophysiology. 2024;67:1077–1079

37. Woodard DA, Kim G, Nilsson KR. Risk profiles and outcomes of patients receiving antibacterial cardiovascular implantable electronic device envelopes: A retrospective analysis. World Journal of Cardiology. 2022;14:177–186

38. Ziacchi M, Biffi M, Iacopino S, Di Silvestro M, Marchese P, Miscio F, et al. Reducing infections through cardiac device envelope: Insight from real world data. The reinforce project. Europace. 2023;25

39. Asbeutah AAA, Salem MH, Asbeutah SA, Abu-Assi MA. The role of an antibiotic envelope in the prevention of major cardiac implantable electronic device infections: A systematic review and meta-analysis. Medicine (Baltimore). 2020;99:e20834

40. Ullah W, Nadeem N, Haq S, Thelmo FL, Abdullah HM, Haas DC. Efficacy of antibacterial envelope in prevention of cardiovascular implantable electronic device infections in high-risk patients: A systematic review and meta-analysis. International Journal of Cardiology. 2020;315:51–56

41. Pranata R, Tondas AE, Vania R, Yuniadi Y. Antibiotic envelope is associated with reduction in cardiac implantable electronic devices infections especially for high-power device-systematic review and meta-analysis. J Arrhythm. 2020;36:166–173

42. Krahn AD, Longtin Y, Philippon F, Birnie DH, Manlucu J, Angaran P, et al. Prevention of arrhythmia device infection trial: The padit trial. Journal of the American College of Cardiology. 2018;72:3098–3109

43. Kay G, Eby EL, Brown B, Lyon J, Eggington S, Kumar G, et al. Cost-effectiveness of tyrx absorbable antibacterial envelope for prevention of cardiovascular implantable electronic device infection. J Med Econ. 2018;21:294–300

44. Wilkoff BL, Boriani G, Mittal S, Poole JE, Kennergren C, Corey GR, et al. Cost-effectiveness of an antibacterial envelope for cardiac implantable electronic device infection prevention in the us healthcare system from the wrap-it trial. Circulation: Arrhythmia and Electrophysiology. 2020;13:e008503

45. Boriani G, Kennergren C, Tarakji KG, Wright DJ, Ahmed FZ, McComb JM, et al. Cost-effectiveness analyses of an absorbable antibacterial envelope for use in patients at increased risk of cardiac implantable electronic device infection in germany, italy, and england. Value Health. 2021;24:930–938

46. Frausing MHJP, Johansen JB, Afonso D, Jørgensen OD, Olsen T, Gerdes C, et al. Cost-effectiveness of an antibacterial envelope for infection prevention in patients undergoing cardiac resynchronization therapy reoperations in denmark. EP Europace. 2023;25:euad159

47. Rennert-May E, Raj SR, Leal J, Exner DV, Manns BJ, Chew DS. Economic evaluation of an absorbable antibiotic envelope for prevention of cardiac implantable electronic device infection. Europace. 2021;23:767–774

48. Lee Y, Kim SS, Choi S-M, Bae C-J, Oh T-H, Kim SE, et al. Rifamycin resistance, rpob gene mutation and clinical outcomes of staphylococcus species isolates from prosthetic joint infections in republic of korea. Journal of Global Antimicrobial Resistance. 2022;28:43–48

49. Leehan JD, Nicholson WL. The spectrum of spontaneous rifampin resistance mutations in the bacillus subtilis rpob gene depends on the growth environment. Applied and Environmental Microbiology. 2021;87:e01237–01221

50. Xiao Y-H, Giske CG, Wei Z-Q, Shen P, Heddini A, Li L-J. Epidemiology and characteristics of antimicrobial resistance in china. Drug resistance updates. 2011;14:236–250

51. Mick V, Domínguez MA, Tubau F, Liñares J, Pujol M, Martín R. Molecular characterization of resistance to rifampicin in an emerging hospital-associated methicillin-resistant staphylococcus aureus clone st228, spain. BMC Microbiol. 2010;10:68

52. Bae S, Kim ES, Lee YW, Jung J, Kim MJ, Chong YP, et al. Clinical and microbiological characteristics of rifampicin-resistant mrsa bacteraemia. Journal of Antimicrobial Chemotherapy. 2023;78:531–539

53. Reitzel RA, Rosenblatt J, Gerges BZ, Jarjour A, Fernández-Cruz A, Raad II. The potential for developing new antimicrobial resistance from the use of medical devices containing chlorhexidine, minocycline, rifampicin and their combinations: A systematic review. JAC-Antimicrobial Resistance. 2020;2:dlaa002

54. Sampath LA, Tambe SM, Modak SM. In vitro and in vivo efficacy of catheters impregnated with antiseptics or antibiotics: Evaluation of the risk of bacterial resistance to the antimicrobials in the catheters. Infection Control & Hospital Epidemiology. 2001;22:640–646

55. Tambe S, Sampath L, Modak S. In vitro evaluation of the risk of developing bacterial resistance to antiseptics and antibiotics used in medical devices. Journal of Antimicrobial Chemotherapy. 2001;47:589–598

